# Mobile Phone Survey Estimates of Perinatal Mortality in Malawi: A Comparison of Data from Truncated and Full Pregnancy Histories

**DOI:** 10.1101/2024.07.11.24310265

**Authors:** Georges Reniers, Julio Romero-Prieto, Michael Chasukwa, Funny Muthema, Sarah Walters, Bruno Masquelier, Jethro Banda, Emmanuel Souza, Boniface Dulani

## Abstract

**Objectives:** In many Low- and Middle-Income Countries, perinatal mortality estimates are derived retrospectively from periodically conducted household surveys. Mobile phone surveys offer advantages in terms of cost and ease of implementation. However, their suitability for monitoring perinatal mortality has not been established.

**Methods:** We use data from the Malawi Rapid Mortality Mobile Phone Survey (RaMMPS) to estimate perinatal mortality rates from two versions of the survey instrument: a Full Pregnancy History (FPH) and a shorter Truncated Pregnancy History (TPH). Female respondents of reproductive age were randomly allocated to either of these instruments. The sample was generated through random digit dialling (RDD) with active strata monitoring. Post-stratification weighting was used to correct for sample selection bias, and estimates are reported with bootstrap confidence intervals. We estimated the stillbirth rate as the synthetic cohort probability of a foetal death with 28+ weeks of gestation over all pregnancies reaching the same gestational age. The perinatal and extended perinatal mortality rates were defined as the probabilities of dying between 28 weeks and 7 or 28 days of life, respectively. RaMMPS estimates are compared to the 2015-16 Malawi Demographic and Health Survey, and estimates published by the United Nations Inter-agency Group for Child Mortality Estimation (UN-IGME).

**Results:** TPH and FPH were administered for 2,117 and 2,086 women, respectively. Weighted point estimates of the stillbirth (19.38 deaths per 1,000 pregnancies, 95%-Confidence Interval (CI): 14.03-25.42), perinatal (42.00, 95%-CI: 34.27-50.78), and extended perinatal mortality rates (49.57, 95%-CI: 41.62-59.43) from the FPH instrument are in line with DHS and UN-IGME estimates. In comparison, the stillbirth rate from the TPH instrument is biased upwards. Post-stratification weighting produces a small upwards adjustment in the estimates.

**Conclusion:** MPS are a promising method for collecting perinatal mortality data. The FPH instrument produces more plausible results than the shorter TPH questionnaire where the window of retrospection is restricted.

## Introduction and background

The UN estimated that 1.9 million babies were stillborn in 2021, and 2.3 million children died in the first month of life(1). Regional disparities in the burden of perinatal mortality are large, and recent progress in the reduction of stillbirths and neonatal mortality has been modest in comparison to maternal and post-neonatal mortality. UNICEF labelled this *a neglected tragedy* (2). It is also *an invisible tragedy* because most perinatal deaths, including stillbirths, occur in countries where routine administrative data (e.g., Civil Registration and Vital Statistics or Health Management Information Systems) are insufficiently performant to produce estimates that are useful for monitoring progress towards global targets(3, 4). In these settings, perinatal mortality estimates are derived from periodically conducted household surveys, including the Demographic and Health Surveys (DHS).

Owing to the rapid expansion in mobile phone ownership, mobile phone surveys have become an appealing alternative to traditional household surveys. They can be deployed rapidly and without the need for in-person contact; a key feature that makes them more suitable to field in the context of epidemic outbreaks or other humanitarian crisis situations(5). Whereas mobile phone surveys are increasingly common (6, 7), they have not yet been used for measuring stillbirth and neonatal mortality. This study thus aims to ascertain whether plausible perinatal mortality estimates can be generated from a mobile phone survey, and complements another manuscript wherein we used mobile phone survey data to estimate infant and under-five mortality (8).

There are severable possible methodological pitfalls associated with mortality estimation from mobile phone surveys, including acceptability, sample selectivity, and data quality concerns (9-13). One element that might affect acceptability and data quality is the duration of the interview in the sense that longer interviews are more susceptible to interruptions and the respondent’s loss of concentration. The empirical evidence for the latter is not very strong(14), but as long as data quality can be upheld, short duration interviews are desirable for the mere reason that they reduce the burden on the respondent and fieldwork operational costs. In this contribution, we use data from the Malawi *Rapid Mortality Mobile Phone Survey* (RaMMPS) where women of reproductive age were randomly administered one of two different versions of a questionnaire module designed to measure stillbirth and neonatal mortality. One of the criteria on which they differed was the length of the module.

The *Full Pregnancy History* (FPH) questionnaire was adapted from the model DHS questionnaire that was introduced in 2020 (DHS round VIII), after it was established that it was better suited to identify stillbirths than Full Birth Histories with a reproductive calendar (DHS round IV) and supplementary questions for identifying non-live births (DHS round VII)(15, 16). The FPH instrument elicits information about all pregnancies in chronological order, starting with the first pregnancy. We compare the FPH estimates of perinatal mortality with those from a *Truncated Pregnancy History* (TPH) instrument, whereby the data is collected in reverse chronological order until an a-priori defined date is reached. This questionnaire was modelled after truncated birth histories collected in Malaria Indicator Surveys (MIS). Early methodological work evaluating instruments and the order wherein birth or pregnancy histories are to be collected were not always conclusive, but a recent comparison of truncated versus full birth histories suggests that the former produces estimates of child mortality that are biased downwards (17-20).

In the following sections, we describe the data and estimation procedures, and compare the Malawi RaMMPS estimates of the stillbirth and perinatal mortality rates from both questionnaires with estimates from the 2015-16 Malawi DHS and the UN Inter-agency Group for Child Mortality Estimation (UN IGME)(21, 22).

## Data and Methods

### Data and survey instruments

We use data from the national Malawi RaMMPS conducted between 24 January 2022 and 28 July 2023. The fieldwork for this study was coordinated by the Institute of Public Opinion and Research (IPOR, https://www.ipormw.org/) in Zomba, Malawi. The sample for the Malawi RaMMPS was generated via (screened) Random Digit Dialling (RDD) without replacement. Using the mobile phone numbering structure in Malawi, a set of random numbers was generated by Sample Solutions (https://sample.solutions/), and verified against the *Home Location Register* (HLR), which is a database of registered (including pre-paid) numbers on the GSM network. The screening using the HLR identifies the bulk of numbers that are not in use. Thereafter, trained enumerators conducted Computer Assisted Telephone Interviews (CATI) with active strata monitoring(23). Strata were a-priori defined in terms of broad age groups (18-49 and 50-64), sex, region (North, Central, and South), and residential setting (urban/rural). Quotas for each stratum were derived from the 2018 census(24). Once a stratum was filled, respondents with these attributes were no longer eligible to participate in the study. Fieldwork was divided into four blocks of 4 to 5 months each, and quotas were re-set at the beginning of each fieldwork block. Minors below the age of 18 were not interviewed to ensure that all respondents could consent to the interviews themselves. Respondents received 1,200 Malawian Kwacha (∼1.5 USD in 2021) in airtime as compensation for completing the interview. Enumerators worked from their homes, and a random sample of interviews were recorded (with consent) for quality control purposes. A fieldwork supervisor also conducted follow-up calls with respondents who completed the interview.

In comparison to other sub-Saharan African countries, mobile phone ownership in Malawi is relatively low. In 2021, the number of issued SIM cards per 100 individuals in Malawi was estimated at 60% and falls short of the sub-Saharan African average where mobile phone penetration was 93%(25). Mobile phone ownership is particularly low in rural areas, where the gender gap in ownership is also more pronounced. According to the 2015-’16 Malawi DHS, 73.8 and 63.9% of men and women in urban areas owned a mobile phone. In rural areas, 47.1 and 25.9% of men and women owned a phone, respectively (26). In addition, a large majority of the Malawian population lives in rural areas (84% according to the 2018 census (24)). Given these imbalances in population distribution and mobile phone ownership, enumerators had difficulty filling the quotas for rural respondents (women in particular), and this reduced the yield of completed CATIs towards the end of each fieldwork block when the quotas for the easiest to reach respondents had been filled. To alleviate this, we fielded an Interactive Voice Response (IVR) survey to identify rural respondents, and the details are described elsewhere(27).

Starting in the second fieldwork block (26 May 2022), consenting female respondents aged 18-49 were randomly allocated to the TPH or FPH set of questions (supporting information, SI-1). The FPH questions were modelled on the instrument that was used in round VIII of DHS, and solicited information on pregnancy dates, pregnancy outcomes, time of gestation, and the survival status of children(16). As in the DHS, this detailed reproductive history was preceded by a summary pregnancy history, to determine the total number of pregnancies to each woman.

TPH left censor the pregnancies with an end date more than seven years before the interview. Unlike FPH, TPH were recorded in reverse chronological order, an approach that was adapted from the *Malaria Indicator Surveys* (MIS). In order to keep the instrument as short as possible, the TPH instrument did not contain a set of summary pregnancy history questions. The TPH and FPH questionnaire modules used the same set of questions to collect information on the day of birth or pregnancy termination, as well as the gestational age.

For the purposes of the analyses presented here, the pregnancy history data were administratively (left) censored on 1 January 2014. Results with a left censoring date of 1 January 2016 are included as supporting information (SI-3).

### Post-stratification weighting

Mortality estimates from MPS may be affected by selection bias because mobile phone ownership –and possibly also respondent consent– is correlated with respondent characteristics that have a bearing on mortality. The imposition of quotas for the a-priori defined strata only partially alleviates this problem because these are limited to key demographic (age group and sex) and geographic (region and urban/rural place of residence) attributes. It is more challenging to impose quotas on educational background or wealth in a sample constituted via RDD because little or no a-priori information is available on these attributes. We therefore resorted to post-stratification to ensure that the RaMMPS sample is representative of the entire population in terms of a broader number of attributes, including education, household size and household wealth.

Post-stratification weights were estimated by Iterative Proportional Fitting–also known as raking; a method that is regularly used in mobile phone surveys (28). For each fieldwork block, marginal distributions of the RaMMPS data were matched to the female population aged 18-49 in the household roster of the 2015-16 Malawi DHS, which was the most recent nationally representative survey available at the time of writing. Weights were computed and applied for the following attributes: (*i)* age group (18-29, 30-39, or 40-49); (*ii)* urban versus rural place of residence; (*iii)* region (northern, central, or southern); (*iv)* educational attainment (incomplete primary or less, completed primary and incomplete secondary, completed secondary or higher); (*v)* household size (1-4, 5-8, or 9+); and (*vi)* an indicator variable for household-level access to a source of electricity. Post-stratification weights ranged from 0.02 to 12.54 with a mean value of 1.0. Untrimmed weights are used in the manuscript, as this is sometimes recommended for small samples (29, 30). Estimates with trimmed weights are included as supplementary material (SI-3b).

### Stillbirth and perinatal mortality rate estimation

We calculated the–late–stillbirth rate from the RaMMPS data as the synthetic cohort probability of a pregnancy loss with at least 28 weeks of gestation, *q*(28*w, Birth*), and is reported as the number of stillbirths per 1,000 live and stillbirths combined. The perinatal mortality rate is defined analogously but expands the exposure time to the first week of life: *q*(28*w*, 7*d*). For analytical purposes, we also present the extended perinatal mortality rate, covering the first 28 days of life: *q*(28*w*, 28*d*). The latter circumvents the problem of heaping at 7 days and allows us to compare RaMMPS with UN-IGME estimates. UN-IGME estimates are available for the stillbirth rate and the Neonatal Mortality Rate (NMR), but no estimates are published for the probability of dying during the first week of life (i.e., the Early Neonatal Mortality Rate, ENMR). UN-IGME estimates for the perinatal mortality rate are computed by the authors, assuming a log-normal distribution of these indicators.

In order to facilitate direct comparisons with the RaMMPS data, the DHS sample was restricted to women aged 18-49. Further, we used the DHS day of birth–as reported by the mother–and the approximate date of termination (month and year) as informed by the reproductive calendar. UN-IGME estimates are reported for all women of reproductive age, including adolescents aged 15-17, not included in the RaMMPS data.

Confidence intervals (CIs) for RaMMPS estimates were computed via nonparametric bootstrapping, resampling the total number of interviews 1,000 times with replacement. For each sample, probabilities of selection were proportional to the post-stratification weights. The 50^th^ percentile is reported as the central tendency of these distributions; and the 2.5^th^ and 97.5^th^ percentiles were used to report 95% CIs and to test for statistical significance. Confidence intervals for the DHS data are produced using a similar procedure. UN-IGME estimates are reported with 90% confidence bounds.

## Results

Out of the 56,072 mobile phone numbers that were tried, a RaMMPS CATI interview was completed with 13,800 respondents (men and women aged 18-64). Response and refusal rates were 26.55 and 10.42 percent, respectively. These are defined as the number of completed interviews or refusals over the number of respondents who either met the inclusion criteria, or, whose eligibility for inclusion in the study could not be established. As described elsewhere, this response rate is a lower bound estimate because cases with unknown eligibility are included in the denominator(31). The analyses in the remainder of this manuscript are restricted to 4,203 interviews with women aged 18-49. About half of these women were administered a TPH and about half received the FPH instrument (SI-2). The median duration to administer the FPH instrument (including summary pregnancy history questions) was 3.20 minutes (Q1-Q3: 0.80-5.20). The median duration to administer the TPH instrument (not including summary pregnancy history questions) was 2.11 minutes (Q1-Q3: 0.38-3.10).

Table 1 provides the individual and household attributes of the RaMMPS respondents in the two pregnancy history modules and the DHS reference dataset. For the RaMMPS data, we present estimates before and after post-stratification weighting. DHS estimates are given for all female respondents aged 18-49 and for the subgroup of women who own a mobile phone. The DHS data confirm that mobile phone owners are more frequently urban, better educated and more often have access to electricity. This is also reflected in the distribution of these attributes in the unweighted RaMMPS samples (columns 2 and 4). Malawi RaMMPS respondents also appear to come from slightly larger households, and that may be due to the fact that larger households have a greater likelihood of being sampled via RDD, or, that estimated household sizes are biased upwards in RaMMPS. After weighting (columns 1 and 3), the imbalance in the RaMMPS data is largely rectified, and the marginal distribution of these background characteristics matches that in the DHS sample for all women (column 6).

**Table 1:** Background characteristics of female respondents aged 18-49 in RaMMPS (weighted and unweighted) and the 2015-16 Malawi DHS (all women and the subset of mobile phone owners)

| Attributes/Instrument & method : | TPH |  | FPH |  | DHS VII |  |
| --- | --- | --- | --- | --- | --- | --- |
|  | *weighted* | *unweighted* | *weighted* | *unweighted* | *all* | *m. owner* |
| <sup>A</sup> Place of residence: urban | 18.80<br>[17.24, 20.50] | 37.13<br>[35.10, 39.13] | 17.98<br>[16.20, 19.56] | 37.78<br>[35.71, 39.96] | 18.40<br>[18.11, 18.69] | 35.50<sup>A</sup><br>[34.86, 36.10] |
| <sup>B</sup> Region: North | 12.28<br>[10.94, 13.67] | 16.96<br>[15.40, 18.54] | 10.83<br>[9.49, 12.22] | 16.49<br>[14.91, 18.17] | 11.68<br>[11.55, 11.83] | 15.04<sup>B</sup><br>[14.76, 15.34] |
| <sup>C</sup> Central | 40.58<br>[38.55, 42.75] | 36.47<br>[34.48, 38.47] | 44.10<br>[41.88, 46.48] | 37.15<br>[35.19, 39.21] | 42.84<br>[42.58, 43.10] | 41.04<sup>C</sup><br>[40.48, 41.62] |
| <sup>D</sup> South | 47.05<br>[44.92, 49.22] | 46.53<br>[44.50, 48.72] | 45.01<br>[42.67, 47.17] | 46.31<br>[44.22, 48.39] | 45.48<br>[45.23, 45.73] | 43.91<sup>D</sup><br>[43.42, 44.39] |
| <sup>E</sup> Education: less than complete primary | 64.90<br>[62.73, 66.89] | 14.64<br>[13.18, 16.20] | 66.01<br>[63.81, 67.86] | 14.33<br>[12.90, 15.82] | 64.65<br>[63.96, 65.37] | 41.78<sup>E</sup><br>[40.50, 43.19] |
| <sup>F</sup> incomplete secondary | 24.61<br>[22.79, 26.22] | 29.90<br>[28.06, 31.88] | 24.35<br>[22.58, 26.13] | 30.73<br>[28.62, 32.89] | 24.74<br>[24.07, 25.39] | 33.27<sup>F</sup><br>[32.02, 34.63] |
| <sup>G</sup> complete secondary or more | 10.53<br>[9.21, 11.81] | 55.41<br>[53.38, 57.44] | 9.68<br>[8.49, 10.93] | 54.89<br>[52.76, 57.09] | 10.61<br>[10.11, 11.09] | 24.96<sup>G</sup><br>[23.68, 26.10] |
| <sup>H</sup> Household size: 1-4 | 37.60<br>[35.55, 39.68] | 35.47<br>[33.54, 37.55] | 42.52<br>[40.41, 44.85] | 38.78<br>[36.67, 40.94] | 40.21<br>[39.43, 41.09] | 41.42<sup>H</sup><br>[39.88, 42.81] |
| <sup>I</sup> 5-8 | 55.13<br>[52.93, 57.13] | 54.42<br>[52.24, 56.47] | 51.05<br>[48.66, 53.19] | 52.16<br>[49.98, 54.48] | 52.85<br>[51.92, 53.66] | 51.82<sup>I</sup><br>[50.41, 53.40] |
| <sup>J</sup> 9+ | 7.27<br>[6.24, 8.46] | 10.01<br>[8.83, 11.38] | 6.42<br>[5.39, 7.50] | 9.06<br>[7.81, 10.26] | 6.92<br>[6.45, 7.44] | 6.78<sup>J</sup><br>[5.98, 7.61] |
| <sup>K</sup> Electricity: access | 13.60<br>[12.23, 15.21] | 52.10<br>[49.98, 54.18] | 12.94<br>[11.48, 14.29] | 53.55<br>[51.37, 55.80] | 13.74<br>[13.22, 14.25] | 30.42<sup>K</sup><br>[29.16, 31.70] |
| <sup>L</sup> Observations | 2117 | 2117 | 2086 | 2086 | 21392 | 8263<sup>L</sup> |

Notes: Both RaMMPS and DHS estimates are reported with bootstrapped 95%-percentile CIs and report the median value of the bootstrap distribution as the point estimate. For the DHS, women were resampled within the same cluster. The distribution of background characteristics for the DHS are given for all women aged 18-49 (the reference for computing the post-stratification weights), and the subset of mobile phone owners.

Figure 1 contains two representations of the stillbirth and perinatal mortality indicators. The top row (panels A-C) compares the bootstrapping distribution of the estimates from the two RaMMPS pregnancy history modules along with the UN IGME estimates; and the 2011-16 DHS estimates. The bottom row of Figure 1 (panels D-F) shows the same estimates on a time scale. Figure 1 contains estimates after post-stratification. Both weighted and unweighted estimates are reported in Table 2. As supplementary material (SI-4), we also provide perinatal mortality estimates by the background characteristics that are used for post-stratification weighting. Unless stated differently, we refer to weighted RaMMPS estimates.

**Table 2:** Malawi stillbirth and perinatal mortality rates by source, before and after post-stratification weighting.

| Instrument/post-stratification | Stillbirth [28w,B) |  | Perinatal [28w,7d) |  | Perinatal [28w,28d) |  |
| --- | --- | --- | --- | --- | --- | --- |
|  | *weighted* | *unweighted* | *weighted* | *unweighted* | *weighted* | *unweighted* |
| <sup>A</sup> TPH, 2014.0-2023.6 | 30.25<br>[22.20, 40.07] | 24.47<br>[16.88, 33.71] | 50.47<br>[40.43, 61.39] | 43.59<br>[33.57, 55.86] | 50.55<br>[40.52, 61.47] | 44.27<sup>A</sup><br>[34.25, 56.53] |
| <sup>B</sup> FPH, 2014.0-2023.6 | 19.38<br>[14.03, 25.42] | 16.06<br>[10.67, 22.48] | 42.00<br>[34.27, 50.78] | 40.45<br>[30.36, 50.89] | 49.57<br>[41.62, 59.43] | 44.50<sup>B</sup><br>[34.44, 55.57] |
| <sup>C</sup> DHS VII, 2011.0-2016.0<br>Women 18-49 | 13.60<br>[11.28, 16.06] |  | 33.89<br>[30.49, 37.83] |  | 38.01<br>[34.24, 42.14] | <sup>C</sup> |
| <sup>D</sup> DHS VII, 2011.0-2016.0<br>Women 18-49, mobile owners | 16.52<br>[11.83, 21.85] | 18.49<br>[13.08, 24.64] | 36.26<br>[29.03, 43.71] | 39.80<br>[32.06, 47.99] | 38.92<br>[31.65, 46.29] | 43.05<sup>D</sup><br>[35.55, 51.35] |
| <sup>E</sup> UN IGME, 2019.5 | 16.15<br>[14.44, 18.10] |  |  |  | 35.96<br>[29.43, 45.17] | <sup>E</sup> |
| <sup>F</sup> UN IGME, 2020.5 | 16.00<br>[13.66, 18.68] |  |  |  | 35.39<br>[28.26, 46.12] | <sup>F</sup> |
| <sup>G</sup> UN IGME, 2021.5 | 15.80<br>[13.04, 19.06] |  |  |  | 34.74<br>[26.81, 46.62] | <sup>G</sup> |

Notes: DHS estimates were computed for all women aged 18-49, and for the subset of mobile phone owners before and after post-stratification weighting. Both RaMMPS and DHS estimates are reported with bootstrapped 95%-percentile CIs and report the median value of the bootstrap distribution as the point estimate. UN-IGME estimates of perinatal mortality are computed by the authors from the stillbirth mortality rate and the neonatal mortality rate.

**Figure 1:**
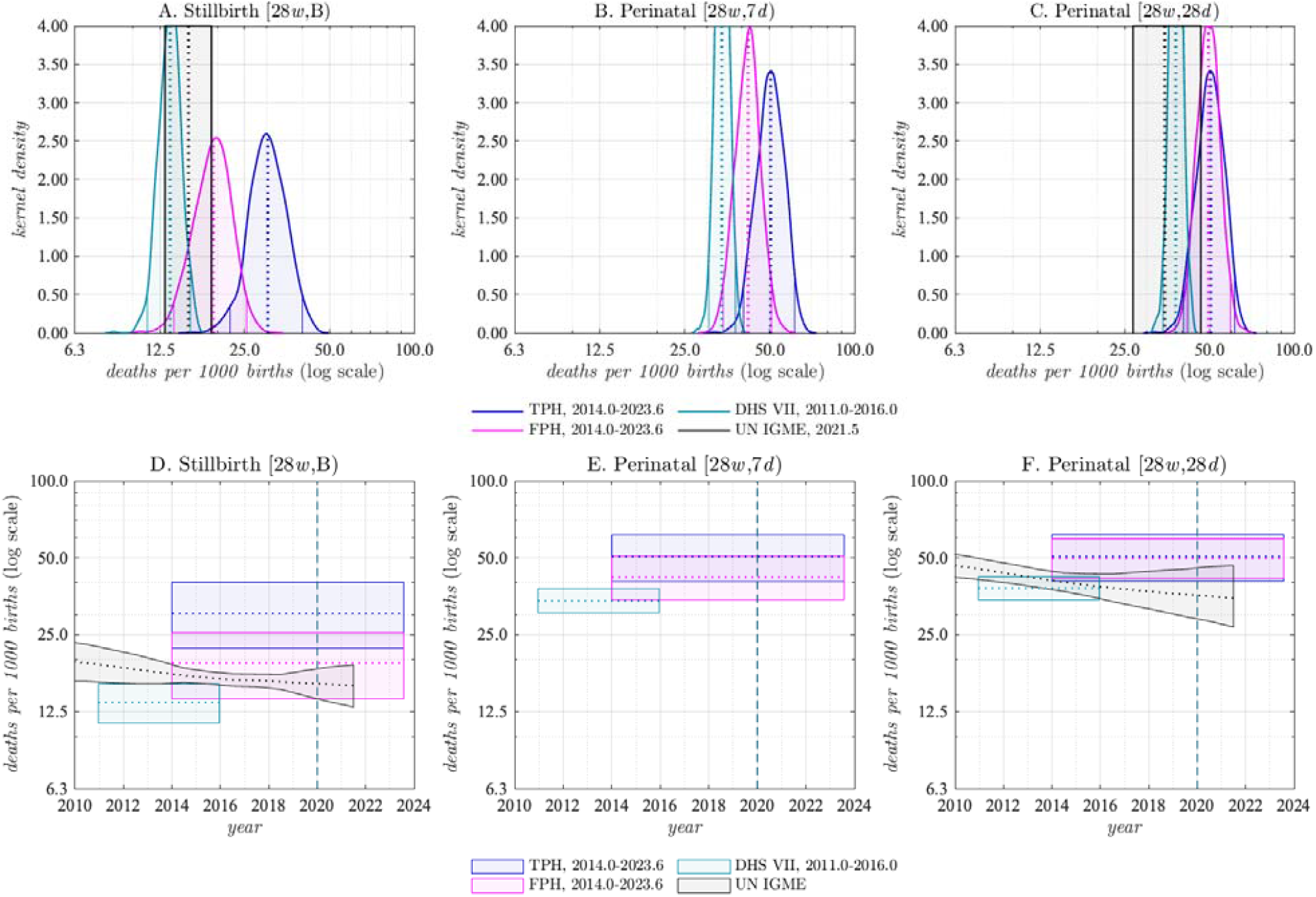
Stillbirth and the perinatal mortality estimates in the Malawi RaMMPS (by survey instrument) compared with UN-IGME and DHS estimates Notes: Panels A-C contain the bootstrap distributions of the RaMMPS and DHS perinatal mortality estimates along with estimates from UN-IGME. In panels D-E, the same estimates are plotted on a time scale and are restricted to the 95%-bootstrap CIs. RaMMPS and DHS estimates pertain to all women aged 18-49. The UN-IGME estimates in panels C and F have been computed by the authors using estimates of stillbirth ratios and the neonatal mortality rate, and assuming a log-normal distribution of these parameters. UN-IGME does not publish separate mortality estimates for the first 7 days of life and it is not possible to compute the UN-IGME equivalent for the indicator in panels B and E. UN-IGME estimates are reported with 90% uncertainty bounds.

**Figure 2:**
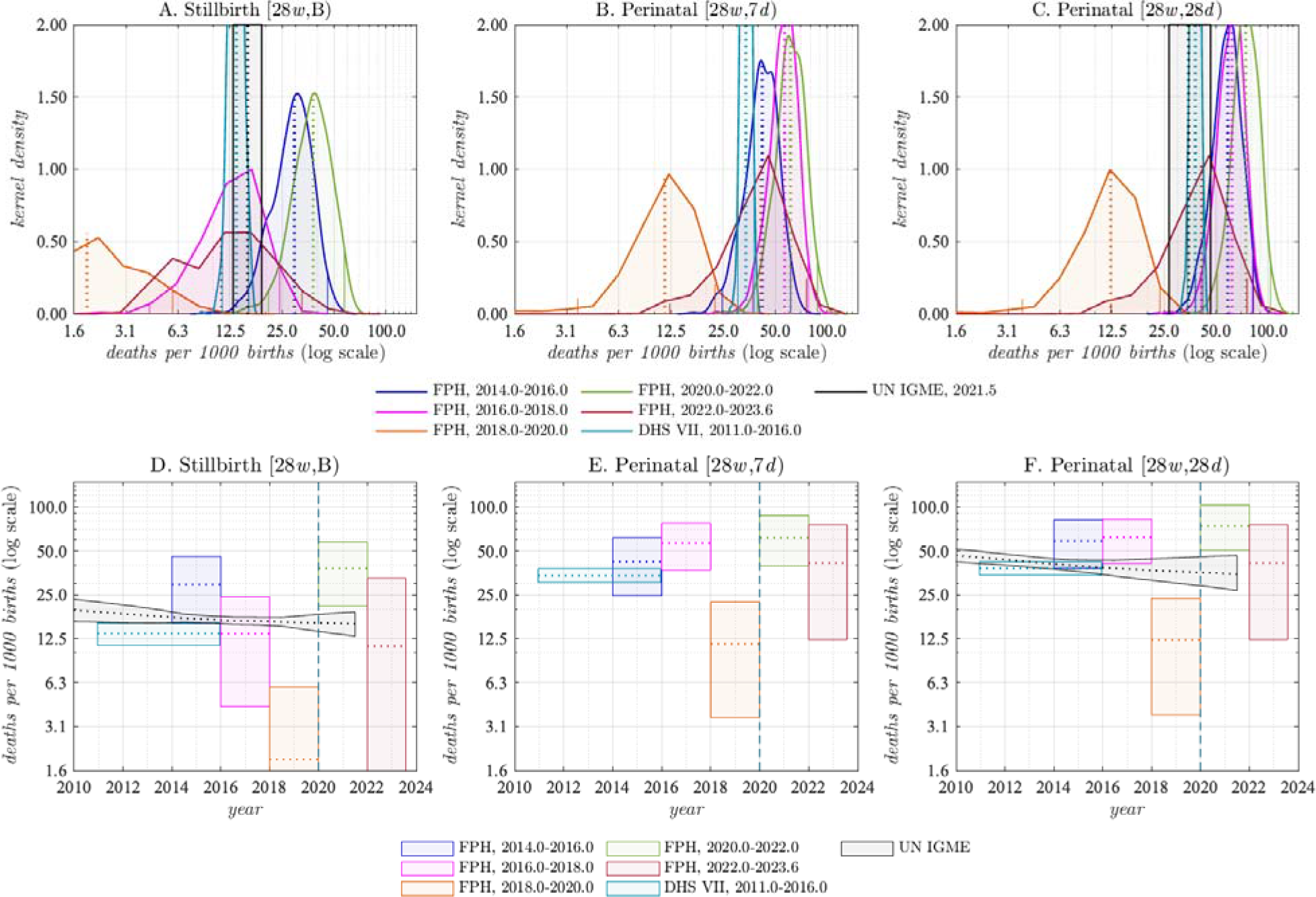
Stillbirth and the perinatal mortality rates in the Malawi RaMMPS by period (FPH instrument only) compared with UN-IGME and DHS estimates Notes: See Figure 1.

The RaMMPS FPH estimate for the stillbirth rate (19.38 per 1,000, 95%-CI: 14.03-25.42) is comparable to that of the UN-IGME estimate for 2019 (16.15, 90%-CI: 14.44-18.10). The DHS estimate is lower, but the difference is not statistically significant. It is also worth noting that the 2015-16 Malawi DHS used the Full Birth History instrument along with a reproductive calendar, and this is now considered to be inferior to a FPH questionnaire for capturing stillbirths(15). The RaMMPS TPH estimate (30.25, 95%-CI: 22.20-40.07) is considerably higher.

The RaMMPS FPH estimate of perinatal mortality (42.00 per 1,000, 95%-CI: 34.27-50.78) and extended perinatal mortality rate (49.57, 95%-CI: 41.62-59.43) exceed estimates from other published sources, but the differences are again not statistically significant. The 2019 UN-IGME estimate for the extended perinatal mortality rate, for example, is 35.96 (90%-CI: 29.43-45.17).

Post-stratification weighting produces a modest upwards adjustment in the RaMMPS perinatal mortality estimates, but they are never significantly different from the unweighted estimates. Application of the post-stratification weighting procedure produces a small downwards correction to the perinatal mortality estimates from the subsample of mobile phone owners from the DHS. Again, the confidence intervals of the weighted, unweighted and full samples overlap.

Figure 3 (SI-5) contains stillbirth and perinatal mortality estimates from the RaMMPS FPH instrument disaggregated by 2-year intervals. Point estimates are indicative of a mortality decline between 2014 to 2020 for each of the indicators that are considered, but they are also suggestive of a temporary mortality reversal in the calendar years corresponding with the COVID-19 outbreak (i.e., 2020-22). The uncertainty around the period-specific FPH estimates is, however, large. Further, the mortality estimates for the period just before and after 2020-22 are very low. While this may result from the stochastic variability in perinatal deaths, it is also possible that there is some displacement of events that artificially inflates the mortality estimates for 2020-22.

One of the complications with evaluating perinatal mortality data and estimates is the absence of a gold standard measurement for most high mortality settings. As done elsewhere, we therefore revert to an evaluation of the age patterns of mortality to ascertain whether stillbirth and early childhood mortality estimates are plausible. In Table 3, this is done in terms of two ratios: (i) the stillbirth to neonatal mortality rate (NMR) ratio, and (ii) the early neonatal (ENMR) to neonatal mortality rate (NMR) ratio.

**Table 3:** Data quality checks: perinatal mortality rate ratios.

| Instrument/post-stratification | Stillbirth [28w,B) | | ENMR - $q(7d)$ | | NMR - $q(28d)$ | | Stillbirth/NMR | | ENMR/NMR | |
| --- | --- | --- | --- | --- | --- | --- | --- | --- | --- | --- |
|  | weighted | unweighted | weighted | unweighted | weighted | unweighted | weighted | unweighted | weighted | unweighted |
| TPH, 2014.0-2023.6 | 30.25<br>[22.20, 40.07] | 24.47<br>[16.88, 33.71] | 20.55<br>[14.73, 27.48] | 19.38<br>[12.98, 27.14] | 20.64<br>[14.80, 27.48] | 20.08<br>[13.55, 27.92] | 1.46<br>[0.95, 2.33] | 1.23<br>[0.75, 2.04] | 1.00<br>[0.97, 1.00] | 0.97<br>[0.89, 1.00] |
| FPH, 2014.0-2023.6 | 19.38<br>[14.03, 25.42] | 16.06<br>[10.67, 22.48] | 23.05<br>[16.69, 29.62] | 24.62<br>[17.40, 33.08] | 30.96<br>[23.63, 38.60] | 28.85<br>[21.05, 37.52] | 0.63<br>[0.41, 0.93] | 0.56<br>[0.34, 0.89] | 0.74<br>[0.64, 0.86] | 0.86<br>[0.75, 0.95] |
| DHS VII, 2011.0-2016.0<br>Women 18-49 | 13.60<br>[11.28, 16.06] |  | 20.62<br>[18.07, 23.58] |  | 24.78<br>[21.75, 27.92] |  | 0.55<br>[0.44, 0.67] |  | 0.83<br>[0.79, 0.87] |  |
| DHS VII, 2011.0-2016.0<br>Women 18-49, mobile owners | 16.52<br>[11.83, 21.85] | 18.49<br>[13.08, 24.64] | 19.89<br>[14.59, 25.74] | 21.69<br>[16.47, 27.76] | 22.68<br>[17.33, 28.64] | 25.07<br>[18.95, 31.35] | 0.73<br>[0.48, 1.08] | 0.74<br>[0.50, 1.08] | 0.88<br>[0.81, 0.94] | 0.87<br>[0.80, 0.93] |

Notes: 95% Bootstrap confidence intervals.

The stillbirth to NMR ratio from the FPH instrument in the Malawi RaMMPS (0.63, 95%-CI: 0.41-0.93) is on par with the 2015-16 Malawi DHS. Median values for the stillbirth to NMR across DHS rounds (all countries) with birth/pregnancy histories range from 0.43 to 0.67(16). Estimates of the ratio from population-based prospectively-collected data in LMICs are somewhat higher: 0.83 (95%-CI: 0.78-.89)(32). The TPH instrument estimate of the stillbirth to NMR ratio (1.46, 95%-CI: 0.95-2.33) is considerably higher than empirical estimates that are reported elsewhere.

The ENMR to NMR ratio from the FPH instrument in the Malawi RaMMPS (0.74, 95%-CI: 0.64-0.86) also compares well to the Malawi DHS estimates included in Table 3. Median estimates of this ratio across DHS rounds ranges from 0.69 to 0.81 (16). Again, the TPH estimate is higher than estimates from other sources.

## Discussion

We have used –for the first time– mobile phone survey data for estimating perinatal mortality via the Truncated and Full Pregnancy History instruments (TPH and FPH). These questionnaires were adapted from those used in face-to-face surveys, and female respondents (aged 18-49) were randomly allocated to either of these two instruments.

The FPH instrument produces point estimates of the stillbirth (19.38, 95%-CI; 14.03-25.42) and (extended) perinatal mortality rates (49.57, 95%-CI: 41.62-59.43) that are comparable to those published by the UN-IGME and the 2015-16 Malawi DHS. The TPH estimate for the stillbirth rate is considerably higher and less plausible. This is corroborated by the data quality checks in terms of the stillbirth to NMR ratio, which is uncharacteristically high for the TPH instrument. The same holds for the fraction of neonatal deaths that occur in the first week (i.e., the ENMR to NMR ratio). In contrast, the extended perinatal mortality rate estimates for both survey instruments are statistically equivalent and also comparable to the DHS and UN-IGME.

The time gained from administering the shorter TPH instrument –amounting to a difference in the median duration of just over one minute– hardly justifies the use of the truncated survey instrument, so our results support the use of FPH as the preferred questionnaire for measuring perinatal mortality in a mobile phone survey. Further, it is worth noting that the time needed to collect pregnancy histories over the phone (typically less than 5 minutes) was considerably shorter than data collection in a face-to-face survey, for which a mean duration of around 10 minutes has been reported (15). Whether this has repercussions for data quality could not be established, but it is certainly an element that requires further consideration. Factors that might contribute to shorter interview durations in a mobile phone survey are the lower fertility rates among mobile phone owners, and differences in the conversational style in a telephone versus an in-person interview.

Selection bias is a systemic problem in mobile phone surveys, and particularly so in circumstances where mobile phone ownership is not universal and possibly correlated with the outcomes of interest. To minimize or circumvent this problem, we have (i) used a quota sample with active strata monitoring and (ii) used post-stratification weights to ensure that our sample represents the population of interest on a number of socio-demographic background characteristics. Because we imposed sampling quotas for urban and rural respondents, the application of post-stratification weights produced a relatively small upwards adjustment in the perinatal mortality estimates only. As argued elsewhere, this approach seems suitable for correcting mobile phone-based mortality estimates, but may be insufficient to recover population estimates for indicators (e.g., contraceptive use or fertility) that are an expression of preferences in addition to one’s socio-demographic attributes (11, 12).

This study was limited by its relatively small sample size. First, this was driven by the difficulty to identify and reach rural women; a problem that may be less pronounced in populations where mobile phone ownership is higher and the gender divide smaller than in Malawi. Second, we introduced the FPH and the randomized comparison of both survey instruments only during the fourth month of fieldwork, following the publication of another study that suggested shortened (a.k.a. *truncated*) instruments tend to produce biased mortality estimates of under-five mortality (17).

Owing to the relatively small sample sizes in this study, it is not possible to draw firm conclusions about mortality differentials over time, or, mortality differentials by the respondent’s background characteristics.

## Conclusion

Mobile phone surveys are a promising tool for collecting perinatal mortality data where birth and death registration is incomplete, and whenever an alternative to an in-person survey is needed. In comparison to the Truncated Pregnancy History (TPH) instrument, Full Pregnancy Histories (FPH) produce more plausible estimates of both mortality levels and age patterns. Because the additional time needed to collect FPH is marginal, we advocate the use of the latter in future mobile phone surveys.

## Data Availability

Malawi RaMMPS data can be requested via email to the corresponding author or. Code to reproduce the results is available from the GitHub repository maintained by JRP (https://github.com/Romero-Prieto/RaMMPS_Malawi)

## Declarations

### Ethics approval and consent to participate

The study protocol was reviewed Ethics Committee of the London School of Hygiene and Tropical Medicine (reference: 26393/RR/24486) and the University of Malawi Research Ethics Committee (reference: P.07/21/76). All participants provided oral consent for the survey, including consent for storing anonymized data in a public repository and consent to audio record the interview.

### Availability of data and materials

Malawi RaMMPS data can be requested via email to the corresponding author or. Code to reproduce the results is available from a GitHub repository (https://github.com/rammps-lshtm/RaMMPS_Malawi_perinatal)

## Authors’ contributions

GR: conceptualization, funding acquisition, fieldwork supervision, methodology, writing. JRP: conceptualization, methodology, formal analysis and visualisation, data curation, writing. MC: fieldwork supervision, review & editing, data curation. FM: fieldwork supervision, review & editing, data curation. SW: methodology, review & editing. BM: funding acquisition, methodology, review & editing. ES: methodology, review & editing. BD: funding acquisition, fieldwork supervision, review & editing, data curation.

All authors read and approved the final version of the manuscript. GR had full access to all of the data in this study and takes complete responsibility for the integrity of the data and the accuracy of the data analysis.

## Acknowledgements

The authors wish to thank that RaMMPS project Scientific Advisory Committee members for inputs on the design and conduct of the project.

