## Supporting Information for "Mobile Phone Survey Estimates of Perinatal Mortality in Malawi: A Comparison of Data from Truncated and Full Pregnancy Histories"

**SI-1 Pregnancy history instruments used in the Malawi RaMMPS**

Starting on 26 May 2022, female respondents of reproductive age were randomly administered a full or truncated pregnancy history questionnaire.

*A. Full Pregnancy History (FPH) questions*

The FPH questionnaire module, was modelled after the standard questionnaire used in the DHS round VIII and starts with question FPH215. This module was preceded by a number of questions about the modalities for the interview (FPH0a-FPH1d), and a set of summary pregnancy history questions (FPH201-FPH211).

| **#** | **Question** | **Response categories** | **Filter** |
| --- | --- | --- | --- |
| FPH0a | Is the respondent still available? | Yes  No | EA3 |
| FPH0b | Please select your (enumerator) gender | Male  Female | PH1a  PH1b |
| FPH1a | INSTRUCTIONS:   - Introduce the topic and ensure that the respondent is sufficiently comfortable discussing births and pregnancies at this time e.g. “I would now like to discuss the births and pregnancies that you have had in your life. Before we do that, I would like to make sure that you are comfortable discussing this topic with me and where you are right now. If you prefer, I can ask a female colleague of mine to call you back at the time that you choose. This follow-up call should not last for more than 5 minutes.” - Discuss whether the respondent can move to a location with greater privacy (if relevant) and record whether the respondent wishes to proceed with the interview.   Do you wish to proceed with the pregnancy history questions at this time? If not, would you prefer to discuss this topic with a female colleague of mine? | Yes  No  Need female interviewer | PH2  PH1c  Reassign |
| FPH1b | INSTRUCTIONS:   - Introduce the topic and reassure yourself that the respondent is sufficiently comfortable discussing births and pregnancies at her current location, e.g., “I would now like to discuss the births and pregnancies that you have had in your life. Before we do that, I would like to make sure that you are in a place where you are comfortable discussing this topic.” - Discuss whether the respondent can move to a location with greater privacy (if need be (and practically feasible) and record whether the respondent wishes to proceed with the interview.   Do you wish to proceed with the pregnancy history questions at this present time? | Yes  No | PH2 |
| FPH1c | On which day do you prefer that we call you back?  INSTRUCTION: A checkbox will allow selection of multiple options. Select as many as apply. | No preference  Monday  Tuesday  Wednesday  Thursday  Friday  Saturday  Refuses callback | END |
| FPH1d | At which time of the day do you prefer that we call you back?  INSTRUCTION: A checkbox will allow selection of multiple options. Select as many as applicable.  For interviews that are rescheduled with the same enumerator, the call outcome should be classified as INCOMPLETE. If the CASE is assigned to another enumerator, the call outcome should be classified as REASSIGNED. | No preference  Before 09:00  9.00-12.00  12.00-14.00  14.00-17.00  17.00-19.00  After 19:00  Refuses callback | END  END  END  END  END  END  END  END |

| FPH201 | Now I would like to ask about all the births you have had during your life.  Have you ever given birth? | Yes  No  Refuse | FPH206  FPH210 |
| --- | --- | --- | --- |
| FPH202 | Do you have any sons or daughters to whom you have given birth who are now living with you? | Yes  No  Refuse | FPH204  FPH204 |
| FPH203a | How many sons live with you? | Number |  |
| FPH203b | How many daughters live with you? | Number |  |
| FPH204 | Do you have any sons or daughters to whom you have given birth who are alive but do not live with you? | Yes  No  Refuse | FPH206  FPH206 |
| FPH205a | How many sons are alive but do not live with you? | Number |  |
| FPH205b | How many daughters are alive but do not live with you? | Number |  |
| FPH206 | Have you ever given birth to a boy or girl who was born alive but later died?  IF NO, PROBE: Any baby who cried, who made any movement, sound, or effort to breathe, or who showed any other signs of life even if for a very short time. | Yes  No  Refuse | FPH208  FPH208 |
| FPH207a | How many boys have died? | Number |  |
| FPH207b | How many girls have died? | Number |  |
| FPH208 | Just to make sure that I have this right: you have had in total [= 203a + 203b + 205a + 205b + 207a + 207b] births during your life. Is that correct?  *INSTRUCTION: Please make sure to go back and change where necessary if the respondent doesn't agree with the Total births* | Yes  No |  |
| FPH210 | Women sometimes have a pregnancy that does not result in a live birth. For example, a pregnancy can end in a miscarriage, an abortion, or the child can be born dead. Have you ever had a pregnancy that did not end in a live birth? | Yes  No | FPH Check 1 |
| FPH211 | How many miscarriages, abortions and stillbirths have you had? | Number |  |
| **FPH CHECK 1:**  Compute the total number of pregnancy outcomes: PO = FPH203a + FPH203b + FPH205a +FPH205b + FPH207a + FPH207b + FPH211  If [PO] = 0: D0  Else if PO > 0: continue | | | |

| Now I would like to ask a few more questions about each of the pregnancies that you had, starting with the first pregnancy that you had | | | |
| --- | --- | --- | --- |
| Define a pregnancy outcome counter POC=1  REPEAT QUESTION 215-230 FOR EACH PREGNANCY | | | |
| FPH215 | Think back to your [first/next] pregnancy. Was that a single or multiple pregnancy? | Single  Multiple  Don’t know | 216  216 |
| FPH215b | Were you pregnant with twins, triplets or more? | Number  Don’t know | MP LOOP |
| REPEAT QUESTIONS FPH216 TO FH230 FOR EACH BABY IN 215b. | | | |
| FPH216 | Was the [First/Second...] baby born alive, born dead, or did you have a miscarriage or abortion? | Alive  Dead  Miscarriage  Abortion  Don’t know | 218  229  229  229 |
| FPH217 | Did the baby cry, move or breathe? | Yes  No  Don’t know | 229  229 |
| FPH218 | Was a name given to this baby? | Yes  No | 219 |
| FPH218b | Write the name | Name |  |
| FPH219 | Is/was [NAME/the baby] a boy or a girl? | Boy  Girl  Don’t know |  |
| FPH220 | What year was (NAME) born?  INSTRUCTION: Please enter -98 if the respondent does not know the year {NAME} was born. | Year |  |
| FPH221 | What month was {NAME} born? | January  February  March  April  May  June  July  August  September  October  November  December  Don’t know  Refuse |  |
| FPH222 | What day of {month – FPH221} was {NAME} born? | NUMERIC (1-31) |  |
| FPH224 | Is name still alive? | Alive  Dead  Refuse | 228 |
| FPH225 | How old was [Name] at a his/her last birthday?  INSTRUCTION: record age in completed years (00 for children below the age of 1 year) | Years | FPH CHECK3 |
| FPH228 | How old was (NAME) when (he/she) died?  INSTRUCTION: record completed days if less than 1 month; completed months if less than two years or completed years if more than two years old. | Days  Months  Years |  |
| FPH CHECK 2:  IF 228 = 12 months OR 1 year à 228a  IF 228 != 12 months OR 1 year à 230 | | | |
| FPH228a | Did (NAME) have his/her first birthday? | Yes  No |  |
| FPH228b | Exactly how many months old was (NAME) when (he/she) died?  RECORD DAYS IF LESS THAN 1 MONTH; MONTHS IF LESS THAN TWO YEARS; OR YEARS. | Days  Months | FPH CHECK 3  FPH CHECK 3 |
| FPH229 | What year did the pregnancy end? | Year  Don’t know | FPH229b |
| FPH229a | Record the year | Year |  |
| FPH229b | What month did the pregnancy end? | January  February  March  April  May  June  July  August  September  October  November  December  Don’t know  Refuse |  |
| FPH230 | How long did this pregnancy last in weeks OR months    INSTRUCTION: record in completed weeks/months | Weeks  Months |  |
| FPH Check 3  MPLoop incomplete à FPH216  MPLoop complete à continue to Update POC + FPH Check 4 | | | |
| Update POC   - Replace POC= POC + 1 if FPH215= [SINGLE/DK] - Replace POC=POC+ [NUMBER] if FPH215 = MULTIPLE   FPH check 4   - If POC < PO à FPH215 (next pregnancy) - If POC ≥ PO à FPH Check 5 | | | |
| FPH231 | Is there any pregnancy that is missing from this list? This could be a pregnancy was not carried to term, a child that is not currently living with you, or a child who passed away. | Yes  No  Don’t know | FPH215 (POC = POC+1)  D0  D0 |

*B. Truncated Pregnancy History (TBH) questions*

The TBH instrument was also preceded by a number of questions about the interview modalities. These are identical to those used in the FPH instrument (FPH0-FPH1) and not repeated here. The TPH instrument loops through pregnancies in reverse chronological order, starting with the most recent pregnancy and ending whenever the last reported pregnancy’s end date was more than 7 years before the survey.

| PH10 | | I would now like to ask you more details about recent pregnancies that you may have had, including pregnancies that did not result in a live birth.  Are you currently pregnant? | | Yes  No  Don’t know  Refuse | PH CHECK 1  PH CHECK 1  PH CHECK 1 |
| --- | --- | --- | --- | --- | --- |
| PH11 | | In how many weeks or months do you expect the baby to come? | | Months  Weeks |  |
| PH CHECK 1:  IF (PH10 = NO or PH10 = DK or PH10 =Refuse): PH12  ELSE IF PH10=YES: PH13 | | | | | |
| PH12 | | Have you ever been pregnant? | | Yes  No  Don’t know  Refuse | PH14a  SSH0  SSH0  SSH0 |
| PH13 | | Have you ever had a pregnancy before the current one? | | Yes  No  Don’t know  Refuse | PH14b  SSH0  SSH0  SSH0 |
| PH14a | | I would now like you to think about the last pregnancy that you have had.  Was that a single or multiple pregnancy? | | Single  Multiple  Don’t know | PH16  PH15  PH16-MP LOOPx1 |
| PH14b | | I would now like you to think about the previous pregnancy that you had before the one we just discussed.  Was that a single or multiple pregnancy? | | Single  Multiple  Don’t know | PH16  PH15  PH16 |
| PH15 | | Were you pregnant with twins, triplets or more? | | Number  Don’t know | MP LOOP x2 |
| PH15a | | How many (twins, triplets, or more?)? | | Number | MP LOOP x N |
| PH16 | | Was the (first/second/third) baby born alive, born dead, or did you have a miscarriage or abortion? | | Alive  Dead  Miscarriage  Abortion  Refuse  Don’t know | PH18  PH25  PH25  PH25 |
| PH17 | | Did the baby cry, move, or breathe? | | Yes  No  Don’t know | PH25  PH25 |
| PH18 | | Was a name was given to this baby? | | Yes  No | PH19 |
| PH18a | | What is the name? | | Name |  |
| PH19 | | Is/Was [NAME/the baby] a boy or a girl? | | Boy  Girl  Don’t know |  |
| PH20 | | In which month was [NAME/the baby] born? | | January  February  March  April  May  June  July  August  September  October November  December  Don’t know |  |
| PH20b | | In which year was [NAME/the baby] born? | | Year |  |
| PH21 | | How long did this pregnancy last in weeks or months?  INSTRUCTION: record in completed weeks/months | | Weeks  Months |  |
| PH22 | | Is [NAME/the baby] still alive? | | ALIVE  DEAD  Refuse | PH24  PH CHECK 2 |
| PH23 | | How old is [NAME/the baby] today?  Hint: {NAME} was born in {birth month}, {birth year}.  INSTRUCTION: record days if less than 1 month; completed months if less than two years or completed years if more than two years old. | | DAYS  MONTHS  YEARS | PH23b  PH23c |
| PH24 | | How old was (NAME) when s/he died?  INSTRUCTION: record completed days if less than 1 month; completed months if less than two years or completed years if more than two years old. | | DAYS  MONTHS  YEARS  Refuse | PH24b  PH24c  PS CHECK 2 |
| PH25 | | In which month did the pregnancy end? | | January  February  March  April  May  June  July  August  September  October  November  December  Don’t know |  |
| PH25a | | In which year did the pregnancy end? | | INTEGER (4 digits) |  |
| PH26 | | How long did this pregnancy last in weeks or months    INSTRUCTION: record in completed weeks/months | | WEEKS  MONTH |  |
| PH CHECK 2:  Multiple pregnancy loop incomplete: PH16  Multiple pregnancy loop complete: PH CHECK 3 | | | | | |
| PH CHECK 3:  Date (interview) – (PH20b or PH25a) ≤ 7 years: PH26loop  Date (interview) – (PH20b or PH25a) > 7 years: PH Check 4 | | | | | |
| PH26 loop | | | Have you ever had a pregnancy before the one we just discussed? | Yes  No  Don’t know  Refuse | NOTE 1  Continue  Continue  Continue |
| PH Check 4:  Thank you for providing all these details about your recent pregnancies. I would now quickly like to review these with you. I have so far recorded that you have had the following pregnancies since 2015:  INSTRUCTION: Please go back to the table to review, modify or proceed.  *Interactive table shows up in SurveyCTO on previous screen and throughout the loops* | | | | | |
| PH27 | Is there any pregnancy that is missing from this list? This could be a pregnancy was not carried to term, a child that is not currently living with you, or a child who passed away. | | | YES  NO  DK | NOTE 2  END  END |
| NOTE1 | INSTRUCTION: Do not read this aloud to the respondent.  Add a new group to discuss the pregnancy.  <add screenshot of what to do> | | |  | Restart loop w/ PH14b |

**SI-2: Pregnancy history data used in the analyses**

Table SI-2: Malawi RaMMPS aggregated pregnancy history data, by survey instrument

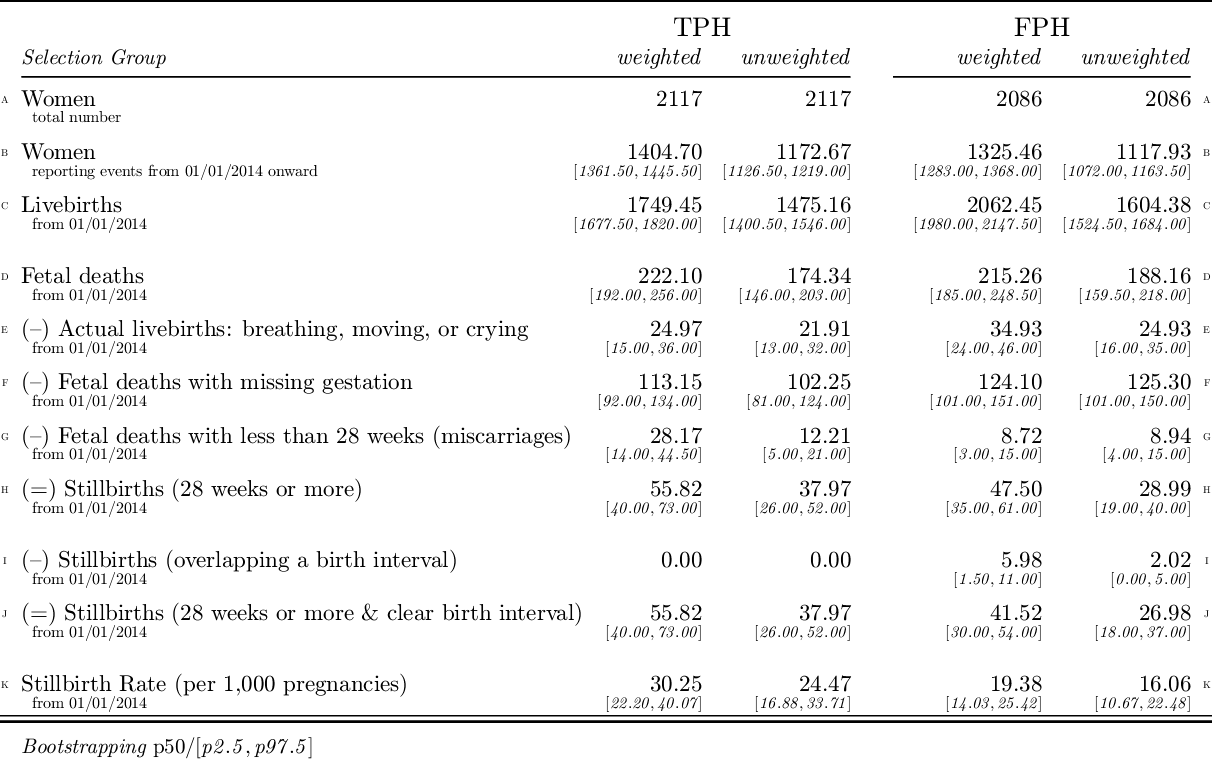

Notes: Reported values correspond to the mean (not median) and the 2.5^th^ and 97.5^th^ percentiles of the Bootstrap distribution. Unweighted counts assume that all women have the same probability of selection, while weighted estimates use the post-stratification weights for resampling purposes.

This table summarizes the pregnancy history data that were used in the analyses. In total, 4,203 women were administered either the full (N=2,086) or truncated (N=2,117) pregnancy history instruments. The analyses reported in this study were restricted to events from the 1 January 2014 onwards, which restricts the effective sample size for the study (row B). Because bootstrapping was used for computing CIs, the totals in rows B-K contain decimals.

The total number of fetal deaths are listed in row D, and the following three rows contain the deductions (e.g., reclassified livebirths, fetal deaths <28 weeks) to arrive at the estimated number of stillbirths. Row I contains the number of reported stillbirths with an implausible pregnancy interval, and these were also dropped from the analyses. The number of stillbirths to compute the stillbirth rate are reported in row J.

**SI-3: Sensitivity analyses**

In this set of sensitivity analyses, estimates from Table 3 in the manuscript are reproduced with an alternative left censoring date (Table SI-3a) and trimmed post-stratification weights (Table SI-3B).

Table SI-3a is useful to evaluate whether transference of events into or out reference window for the TPH instrument affected estimates.

Post-stratification weights ranged from 0.0213 to 12.5398 (with an average value of 1.0000). The median value for the weights weight is 0.3217 and 95% of the weights are less or equal than 5.7347. Estimates where weights were capped at the value for the 95^th^ percentile of the weight distribution are presented in Table SI-3b.

Table SI-3a: Estimates with left censoring on 1 January 2016

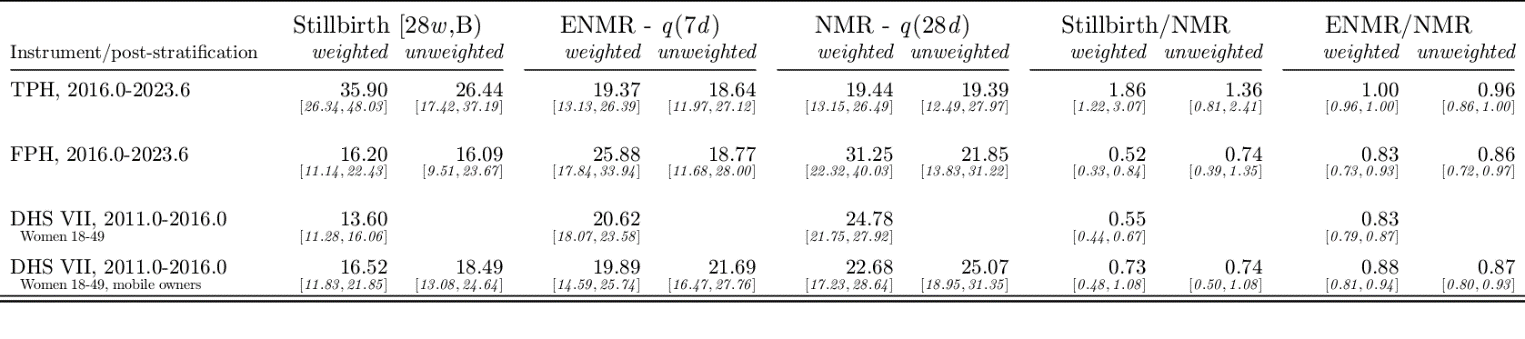

Notes: Estimates are reported with 95% Bootstrap CIs.

Table SI-3b: Estimates with trimmed post-stratification weights

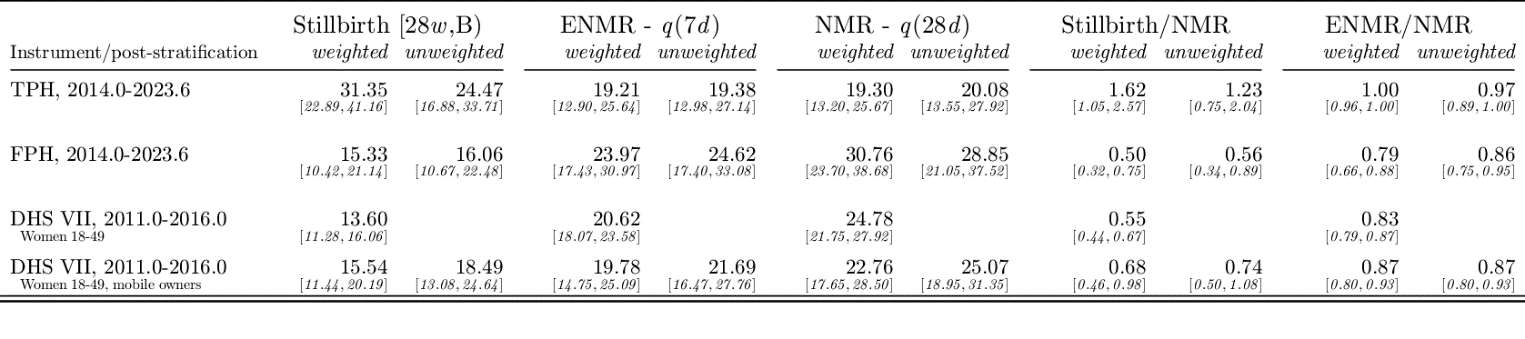

Notes: Estimates are reported with 95% Bootstrap CIs.

**SI-4: Stillbirth and perinatal mortality rates by mother’s attributes**

Tables SI-4a and SI-4b summarize mortality differentials by the background characteristics that are also used as post-stratification weighting variables. As expected, these are suggestive of a social gradient in mortality, but the differences typically fail to reach statistical significance. FPH perinatal mortality estimates are, however, significantly higher for rural areas and for women living in households without electricity (Table SI-4b).

Table SI-4a: Stillbirth rates by mother’s attributes; RaMMPS vs. DHS-VII

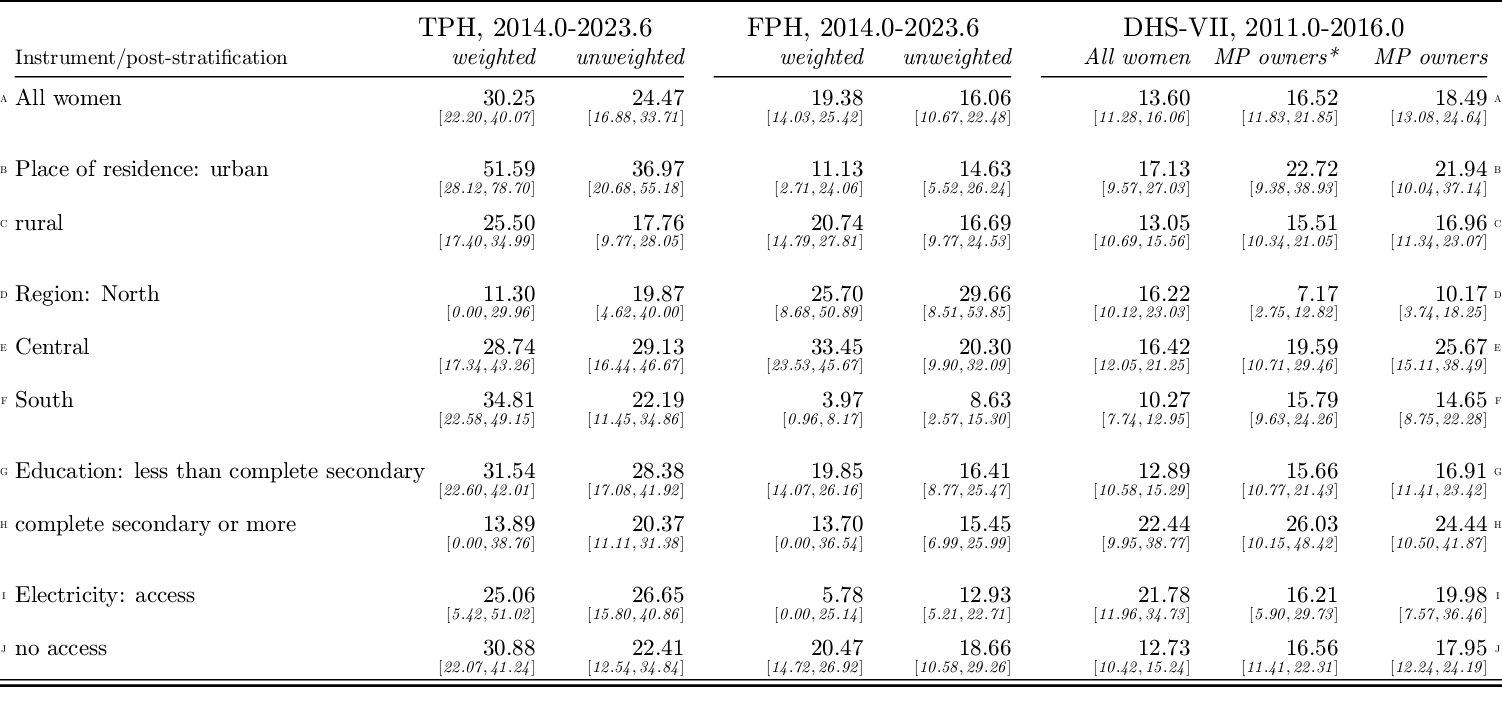

Notes: Estimates are reported with 95% Bootstrap CIs. * weighted estimates using the subsample of mobile phone owners

Table SI-4b: Perinatal mortality rates by mother’s attributes, RaMMPS vs. DHS-VII

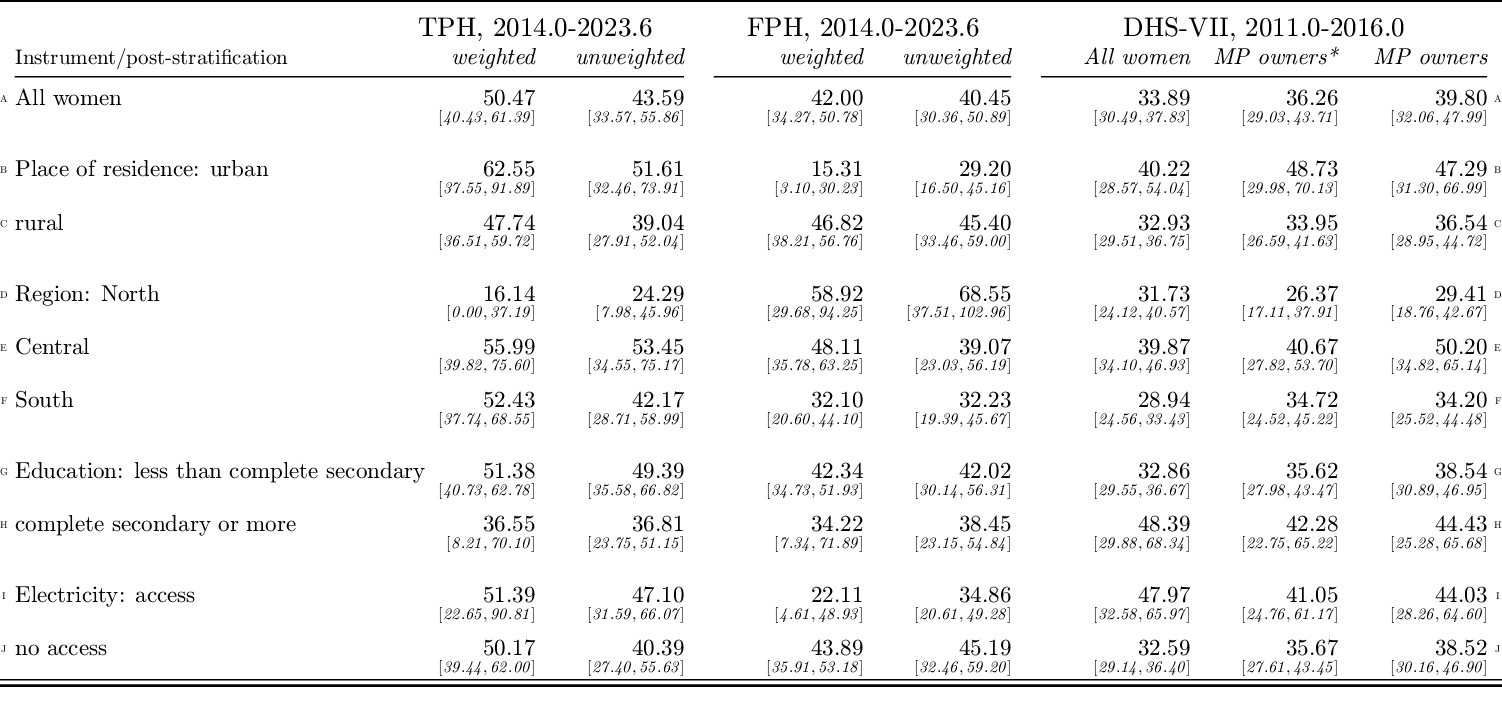

Notes: Estimates are reported with 95% Bootstrap CIs. * weighted estimates using the subsample of mobile phone owners

**SI-5: Trends in stillbirth and perinatal mortality**

Table SI-5: Trends in stillbirth and perinatal mortality estimates

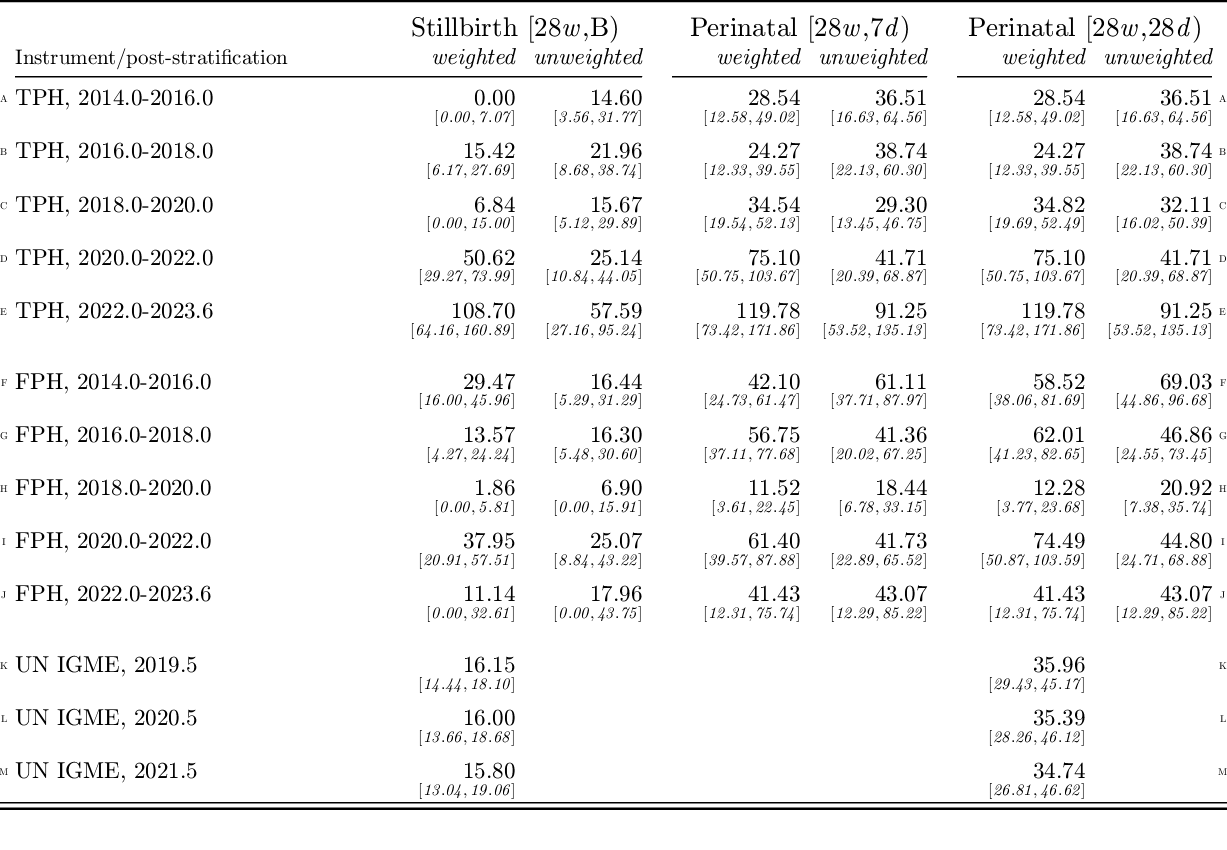

Note: RaMMPS TPH and FPH estimates are reported with 95% Bootstrap CIs. UN-IGME estimates of perinatal mortality are computed by the authors using the stillbirth mortality rate and the neonatal mortality rate. UN-IGME reports 90% confidence bounds
